# Relative Pandemic Severity in Canada and Four Peer Nations During the SARS-CoV-2 Pandemic

**DOI:** 10.1101/2021.03.23.21253873

**Authors:** Amy Peng, Alison E. Simmons, Afia Amoako, Ashleigh R. Tuite, David N. Fisman

## Abstract

**Introduction:** National responses to the SARS-CoV-2 pandemic have been highly variable, which may explain some of the heterogeneity in the pandemic’s health and economic impacts across the world. We sought to explore the effectiveness of the Canadian pandemic response relative to responses in four peer countries with similar political, economic and health systems, and with close historical and cultural ties to Canada (the United States, United Kingdom, France, and Australia) from March 2020 to May 2022.

**Methods:** We used reported age-specific mortality data to generate estimates of pandemic mortality standardized to the Canadian population. Age-specific case fatality, hospitalization, and intensive care admission probabilities for the Canadian province of Ontario were applied to estimated deaths in order to calculate hospitalizations and intensive care admissions averted by the Canadian response. The monetary value of averted hospitalizations was estimated using cost estimates from the Canadian Institute for Health Information. Age-specific quality-adjusted life-years (QALY) lost due to fatality were estimated using published estimates. QALY were monetized using a net expected benefit approach.

**Results:** Relative to the United States, United Kingdom, and France, the Canadian pandemic response was estimated to have averted 94,492, 64,306 and 13,641 deaths respectively, with more than 480,000 hospitalizations averted, and 1 million QALY saved, relative to the United States. A United States pandemic response applied to Canada would have resulted in more than $40 billion in economic losses due to healthcare expenditures and lost QALY; losses relative to the United Kingdom and France would have been $21 billion and $5 billion respectively. By contrast, an Australian pandemic response would have averted over 28,000 additional deaths and averted nearly $9 billion in costs in Canada.

**Conclusions:** Canada outperformed peer countries that aimed for mitigation, rather than elimination, of SARS-CoV-2 in the first two years of the pandemic, likely because of a more stringent public health response to disease transmission. This resulted in substantial numbers of lives saved and economic costs averted. However, comparison with Australia demonstrates that an elimination focus would have allowed Canada to save tens of thousands of lives, and would have saved substantial economic costs.

## Introduction

The global SARS-CoV-2 pandemic has taken a fearsome toll on mortality, life expectancy and population health globally, but not all countries have been impacted equally. The reasons for this heterogeneity are only partly understood; population age structure is a key contributor to SARS-CoV-2 severity (1, 2), but countries with older age distributions (such as Japan) have been less severely affected than high-income peers (3). Japan’s early focus on the airborne nature of SARS-CoV-2, and the widespread acceptance of masking, may also have been important mitigators (3, 4). Marked heterogeneity in severity is seen across countries that have similar age structures, and which were slow to recognize airborne transmission of SARS-CoV-2.

A case in point is the differential severity of the pandemic in Canada and the United States; both are wealthy, federal democracies with advanced medical care systems. In both countries, the COVID-19 pandemic has had a major impact on population health and the economy. The similarities and differences between the two countries’ healthcare systems have made cross-national comparisons an important source of insight into the strengths and weaknesses of their respective health systems (5). During the COVID-19 pandemic, both COVID-19 cases and deaths per capita have been substantially higher in the United States than in Canada (6). By contrast, Australia represents another reasonable comparator for Canada, with similarities in income, culture and governance, but which employed more stringent pandemic control measures, and which has had much lower per capita SARS-CoV-2 pandemic mortality as of May 2022 (7). The United Kingdom and France share ties of economy, culture and history with Canada (as hubs of the British Commonwealth, and La Francophonie, both of which include Canada), and may also represent appropriate comparators.

Debate in the Canadian public sphere around pandemic policy often focusses on whether Canada’s approach to disease control should be more stringent or less stringent. Assuming that differences in outcomes were at least in part driven by policy, rather than the independent actions and choices of individuals, we sought to explore the differences in outcomes that Canada would have experienced over the first two years of the SARS-CoV-2 pandemic, had it followed the path of the United States, the United Kingdom, Australia, or France. We had previously performed such an analysis in March 2021, with comparison restricted to Canada and the United States (6). While our objective was not to perform a formal cost-utility analysis of the Canadian pandemic response relative to responses in these peer nations, the question of costs averted, or excess costs accrued, both through hospitalizations and premature loss of life, is an important one, and we incorporated simple valuations of these quantities into our analysis. These may help inform future cost-utility analyses on this question.

## Methods

We obtained national COVID-19-attributed death estimates from Public Health Agency of Canada, and national health authorities for the United Kingdom, France, the United States and Australia to late April or early May of 2022, as available (7-11). Population estimates were obtained from national census agencies for all countries (12-16). We calculated the number of excess or deficit deaths that would have been expected in Canada under approaches employed in comparator countries using direct standardization (17). Because country death data were reported using slightly different age groupings, we reallocated Canadian deaths to mirror the distribution of SARS-CoV-2 deaths, by two-year age increments, in the province of Ontario (available to January 18, 2022). Standardized mortality ratios for Canada, relative to other countries, were estimated by dividing observed by expected deaths (i.e., the deaths that *would* have occurred with a US-, UK-, France- or Australia-equivalent response). 95% confidence limits for ln(SMR) were calculated by estimating standard errors as (1/A+1/B)^1/2^, where A and B are death counts in each of the two comparator countries, as described in (17).

Observed deaths were subtracted from expected deaths to calculate deaths averted. We divided averted deaths by age-specific case-fatality estimates from Ontario to estimate averted cases. We applied age-specific risks of hospital admission and intensive care admission, derived from Ontario case data, to calculate hospital and intensive care admissions averted. We placed a monetary value on hospitalizations and ICU admissions averted based on Canadian cost estimates generated by the Canadian Institute for Health Information (18). The approach of Briggs et al., modified for the Canadian context by Kirwin et al., was used to estimate quality-adjusted life-years (QALY) lost for deaths occurring in each age group (19, 20). We monetized QALY losses averted by applying a net expected benefit approach, with QALY valued at $30,000 as per Kirwin et al (20). We compared stringency of pandemic responses using the Oxford Government Coronavirus Response Tracker’s Pandemic Stringency Index, obtained from (21). Stringency was plotted against time, and differences in stringency between Canada and other countries were evaluated with the Wiloxon rank-sum test. All input data are publicly available at https://figshare.com/articles/dataset/Estimated_Deaths_Intensive_Care_Admissions_and_Hospitalizations_Averted_in_Canada_during_the_COVID-19_Pandemic/14036549.

## Results

Fewer SARS-CoV-2 related deaths per capita had occurred in Canada than in the United States in all age groups as of May 2022, with SMR significantly less than 1 for all age groups in Canada. A similar pattern was seen when Canada was compared to the United Kingdom, except in children aged 0-14, where there was no significant difference between the two countries (SMR 1.02, 95% CI 0.67 to 1.55). In comparison with France, Canada was found to have experienced significantly fewer deaths per capita in adults aged 40 to 89 years, more deaths than France in those aged 20-29 and 90 and over, and no difference in those under 20 years. When compared to Australia, Canada was found to have had significantly higher SARS-CoV-2 related per capita mortality in all age groups except those aged 10-19, where differences were not significant (SMR 2.24, 95% CI 0.81 to 6.16) (**Table 1**).

**Table 1.**
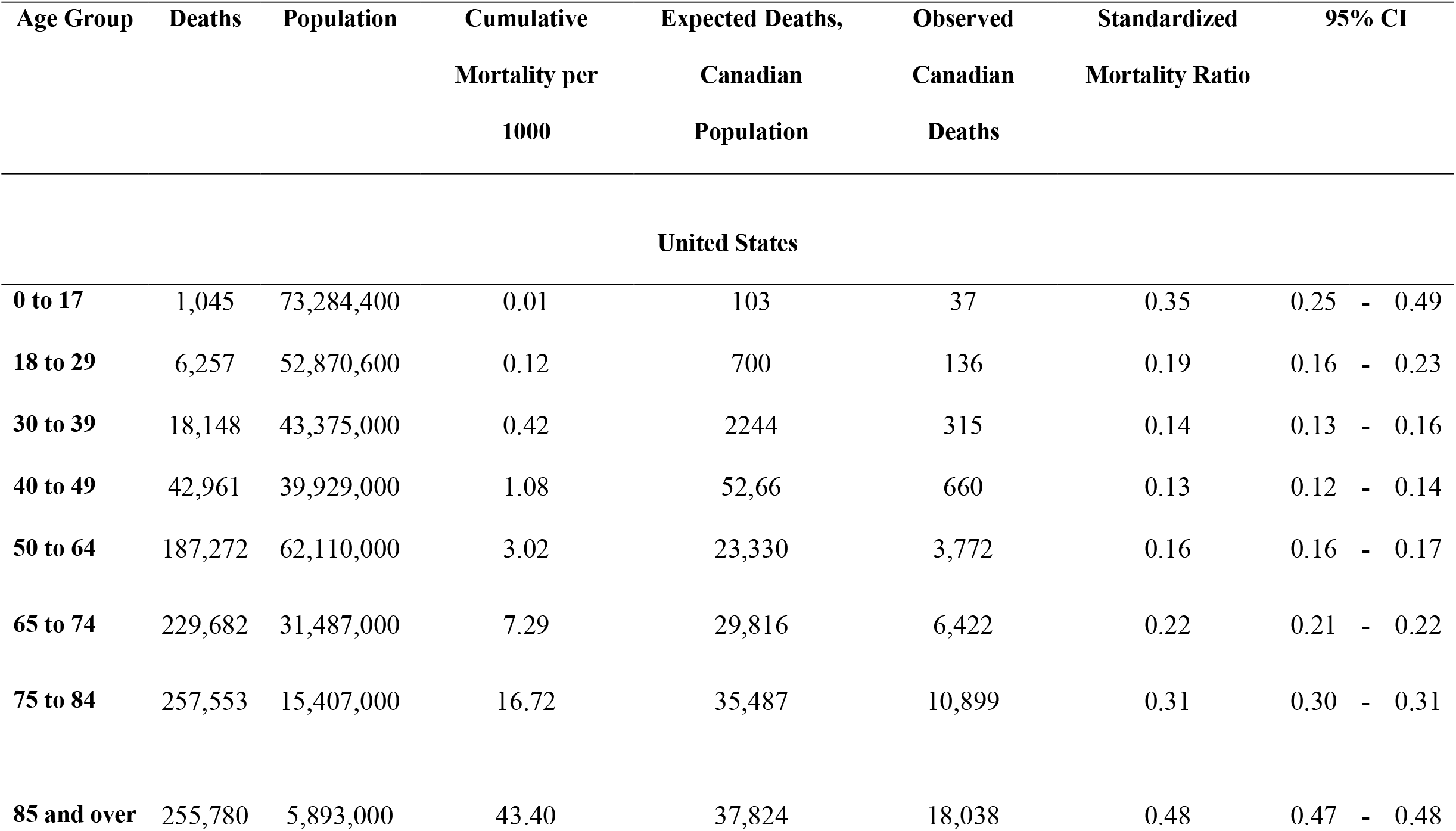
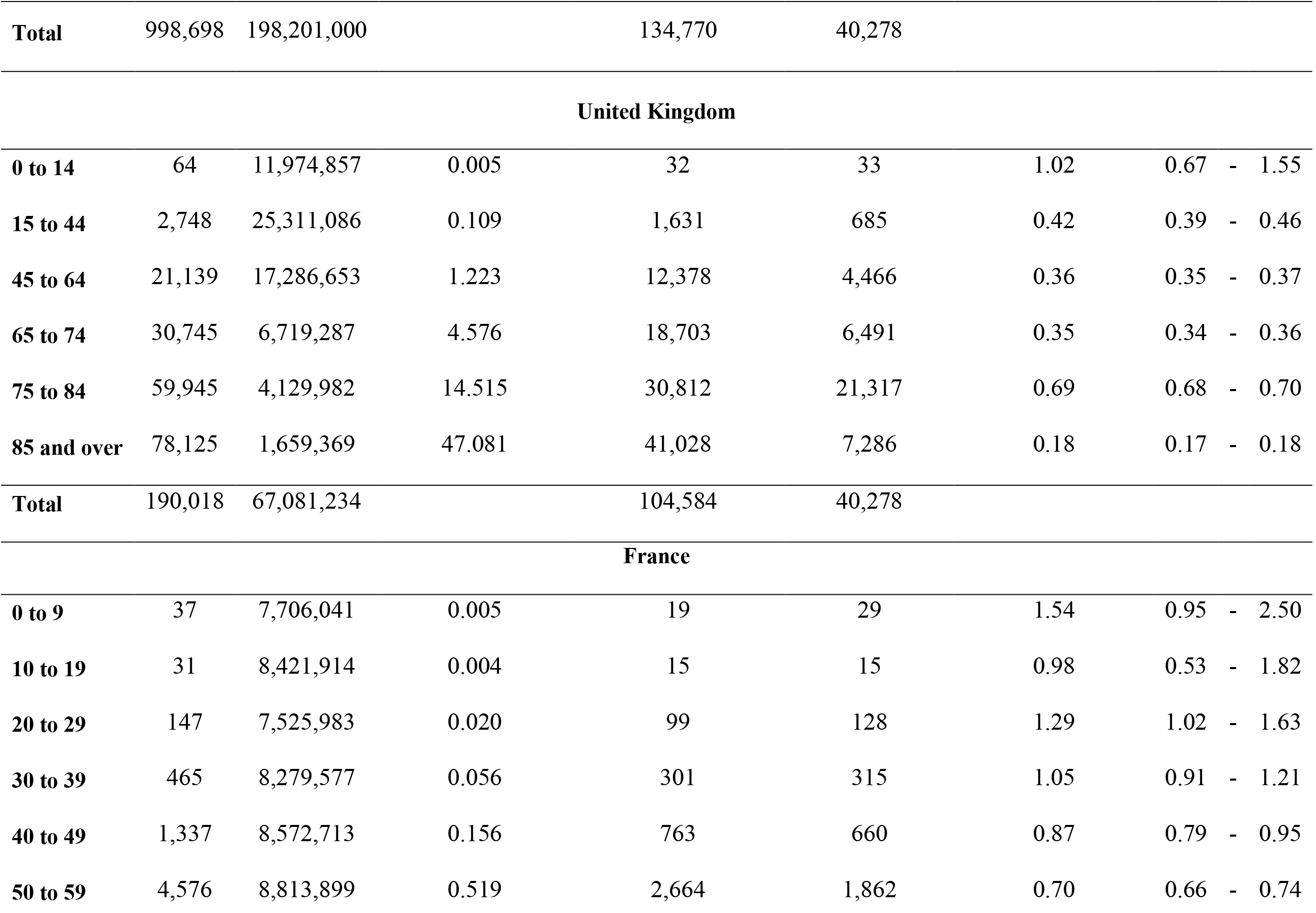
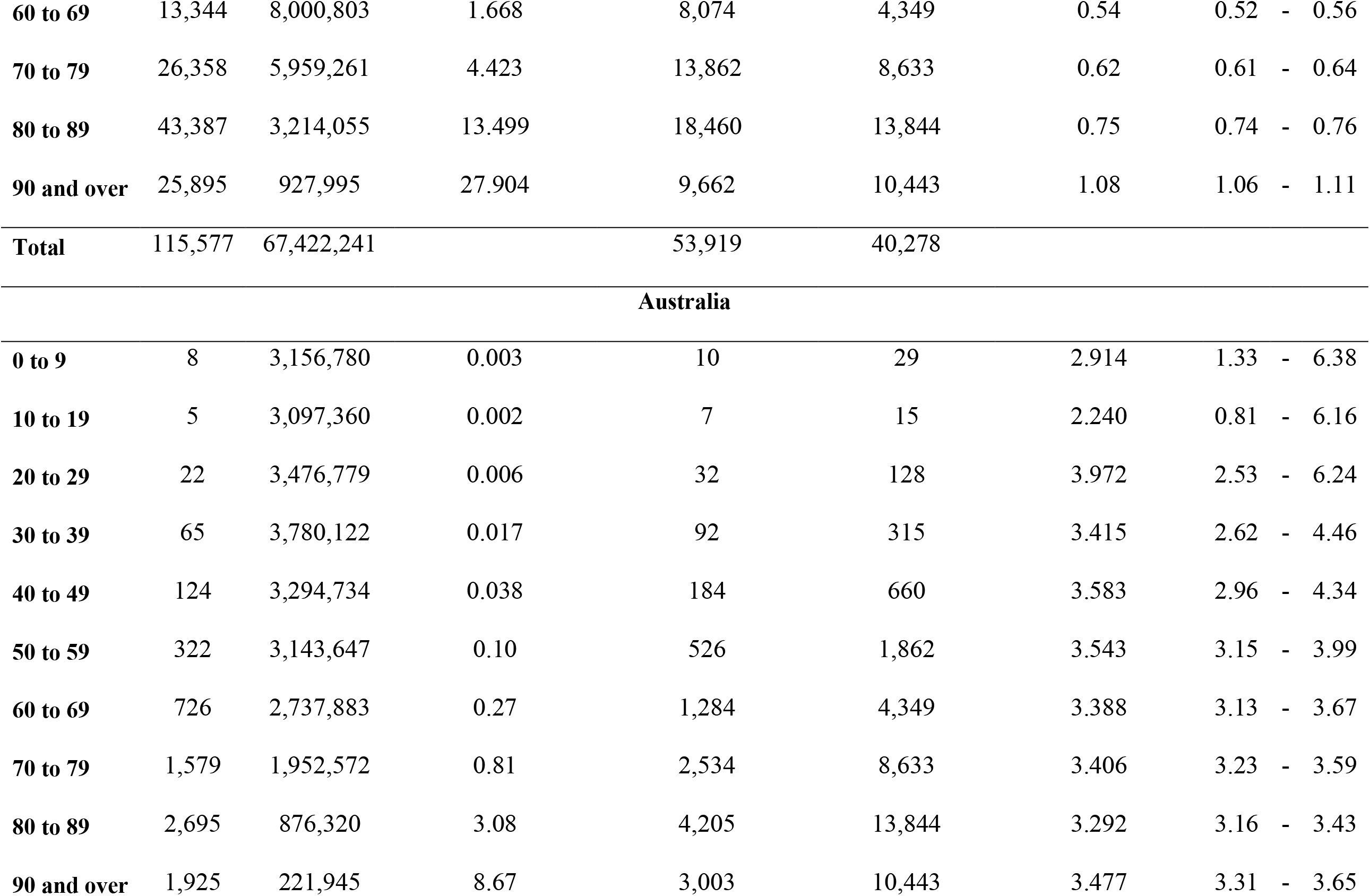
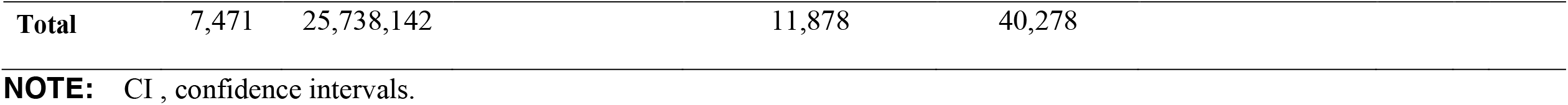
Standardized Mortality Ratios for the First Two Years of the SARS-CoV-2 Pandemic in Peer Countries as Compared to Canada.

When compared to United States, France, and United Kingdom SARS-CoV-2 responses, Canada’s response was estimated to have prevented 94,492 (93,593 to 95,360) deaths, 64,306 (63,394 to 65,189) and 13,641 (12,489 to 14,735) deaths, respectively. By contrast, an Australian response applied to Canada would have saved 28,400 (26,097 to 30,939) lives of the 40,278 that had been lost to SARS-CoV-2 as of May 2022 (**Table 2**). Distributions of deaths by age also differed markedly between the United States and the other countries analyzed. For example, half of deaths in the United States occurred in individuals under the age of 55; in other countries, half of fatalities occurred under the approximate age of 75, with the remainder occurring in those aged 75 and over (**Figure 1**). A similar divergence between the United States response and those in other countries was seen when we applied age-specific QALY losses to death data (**Figure 2**).

**Table 2.**
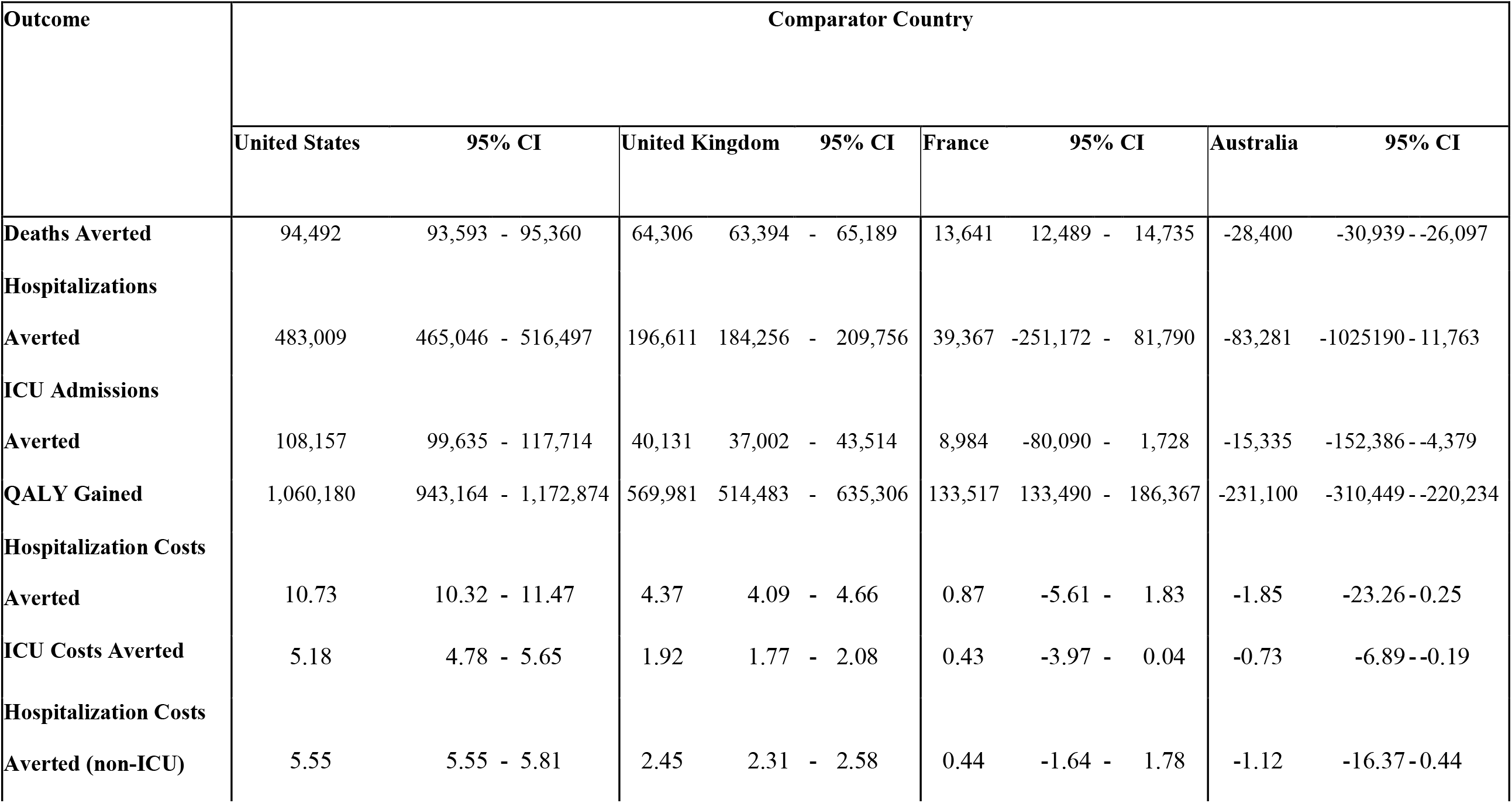
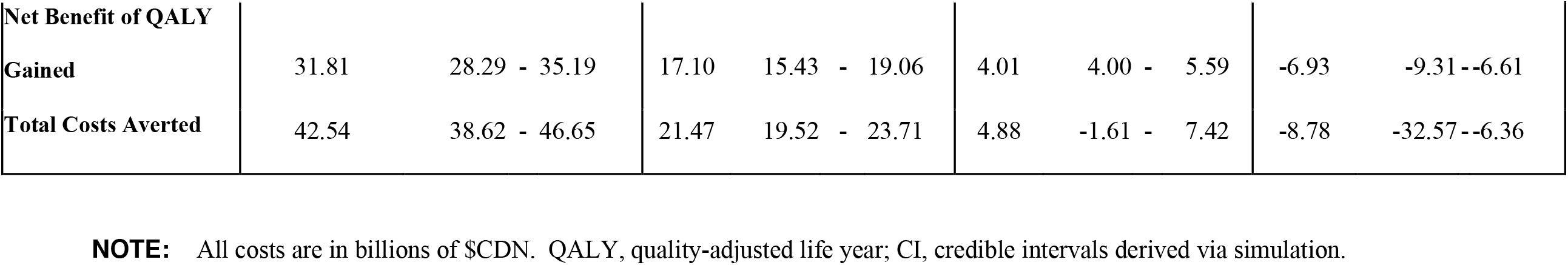
Health Outcomes and Costs Averted in Peer Countries as Compared to Canada.

**Figure 1.**
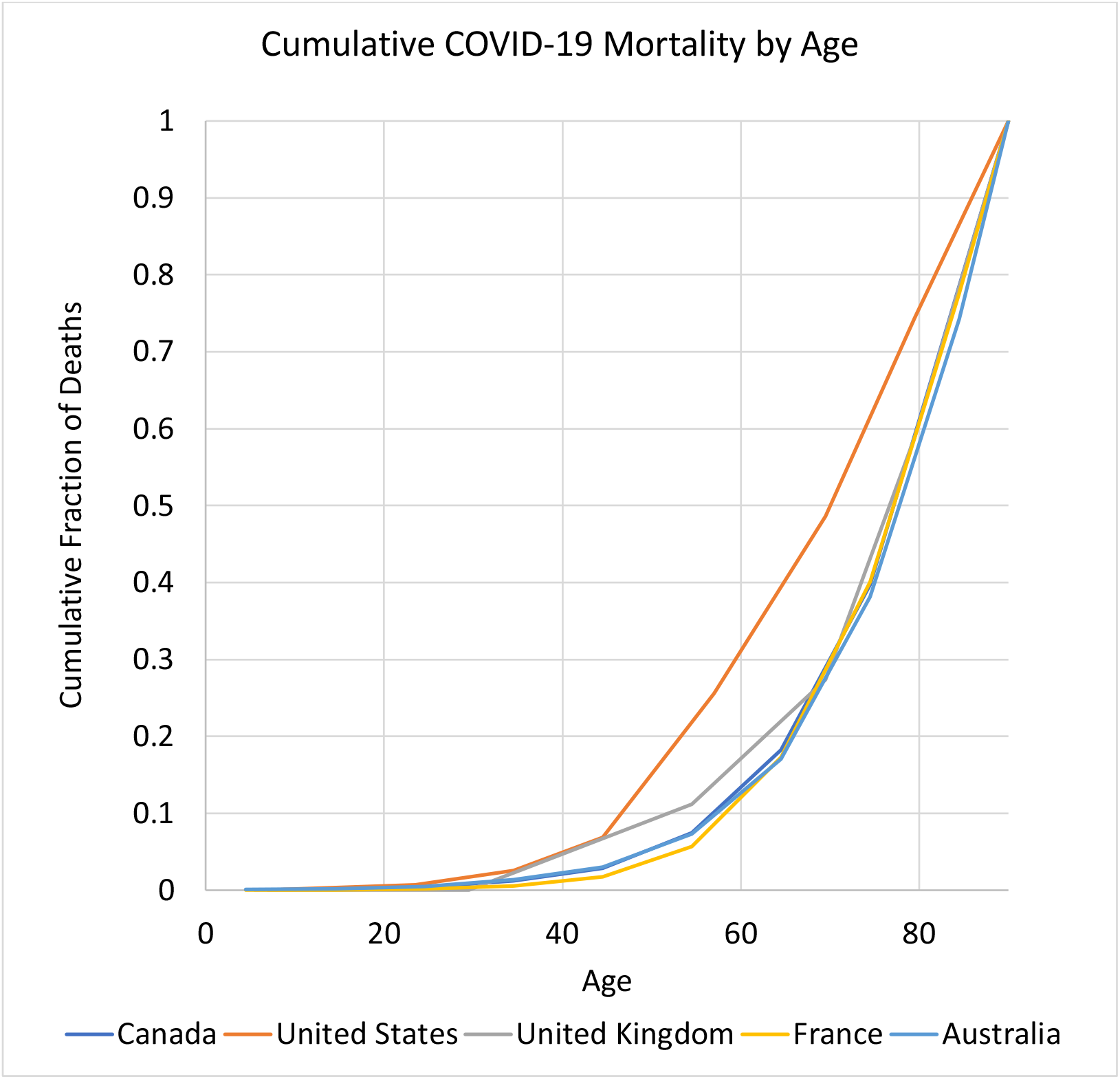
Cumulative Proportion of COVID-19 Attributable Death by Age, March 2020 to May 2022. Ages represent the midpoints of age categories. For the oldest age categories in Canada (80 and over) and the United States (85 and over) we assigned an age of 90 years.

**Figure 2.**
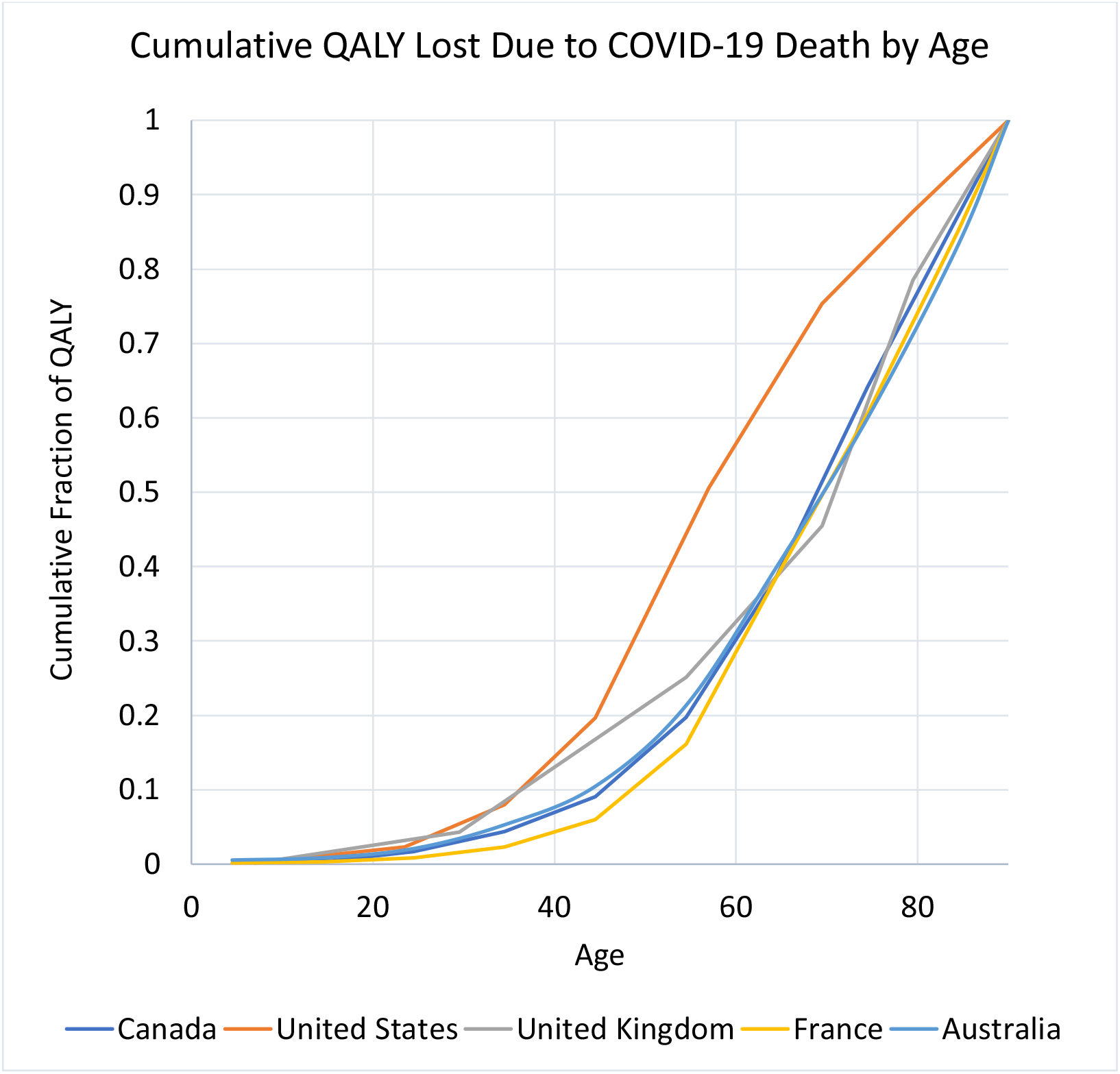
Cumulative Proportion of COVID-19 Attributable Quality-adjusted Life Years (QALY) Loss by Age, March 2020 to May 2022. Ages represent the midpoints of age categories. For the oldest age categories in Canada (80 and over) and the United States (85 and over) we assigned an age of 90 years.

We estimated that Canada’s response saved over 1 million QALYs, nearly 500,000 hospitalizations, and over 100,000 ICU admissions relative to what would have occurred with a response equivalent to that seen in the United States (**Table 2**). The value of QALY losses and hospitalizations averted is estimated to be approximately $43 billion, with $32 billion due to aversion of lost QALY, and the remainder due to averted hospitalizations. The Canadian response also saved QALY and averted hospitalizations and ICU admissions relative to United Kingdom and French responses, though in the case of France credible intervals for estimates of hospitalizations and ICU admissions averted, and costs saved, overlapped zero (cost savings, $4.88 billion, 95% CI -$1.61 to $7.42 billion). When compared to the Australian response, Canada’s response was estimated to have resulted in approximately 230,000 additional QALY lost, over 80,000 excess hospital admissions, and over 15,000 excess ICU admissions as of May 2022, representing a loss of $8.78 ($6.36 to $32.57) billion (**Table 2**). Age-specific estimates of deaths, healthcare utilization and costs averted for each of the four comparator countries are available at https://figshare.com/articles/dataset/Estimated_Deaths_Intensive_Care_Admissions_and_Hospitalizations_Averted_in_Canada_during_the_COVID-19_Pandemic/14036549.

The stringency of the Canadian pandemic response from March 1, 2020 to May 1, 2022, was significantly higher than stringency in the United States, United Kingdom and France, but was also higher than Australian stringency (P < 0.001 for all comparisons) (**Supplementary Table and Figures**).

## Discussion

The cultural similarities and integrated economies of Canada and the United States, which also have very different health systems, has long encouraged comparative research between these two countries (5, 22-24). During the current SARS-CoV-2 pandemic, this type of research has continued, spurred in part by the remarkable difference in the pandemic’s impact on Canada and the United States (25). Here, we demonstrate that application of age-specific US data to Canada results in a far deadlier pandemic, with a more than three-fold increase in total deaths relative to those that had occurred in Canada as of May 2022.

A challenge with this type of comparison is that the United States’ pandemic response has emerged as a global outlier, with SARS-CoV-2 taking a far greater toll in terms of loss of life than in any other high income peer country. The outlier status of the United States has the effect of making Canada-U.S. comparisons predictable in result, perhaps unfairly elevating the effectiveness of the Canadian pandemic response. As such, we also evaluated Canada’s response relative to the United Kingdom and France, the two European countries from whose colonies Canada was created, as well as Australia, which given cultural, political, economic, and historical similarities to Canada, is also a fair comparator.

We find that, as with the United States, application of the United Kingdom’s pandemic response to Canada would have resulted in tens of thousands of additional deaths, as well as billions of dollars in excess economic losses. While Canada appears to have outperformed France as well, differences between these two countries are more modest. Australia emerges as a model of what Canada might have achieved by taking a more aggressive stance on disease control during the first two years of the SARS-CoV-2 pandemic; we estimate that over 75% of Canadian pandemic deaths to date could have been averted through an Australian response, with cost-savings of approximately $10 billion.

Our work complements that of Razak et al., who also found that Canada had outperformed most of its G10 peers (Japan excepted) with respect to pandemic attributable mortality (26). However, the use of standardization, as applied here, allows us to see that the Canadian approach was far more effective than the US and UK approaches in preventing deaths in younger adults, with consequently greater gains in quality-adjusted survival. As public health and government officials in these five countries likely had access to similar information for decision-making, differences in outcomes likely reflected active policy choices. The complexity of the pandemic, and societal responses to it, makes identification of causal factors challenging. Galvani et al. noted that a key difference between Canada and the United States may relate to universal public healthcare in the former (25); however, universal public healthcare is also available in Australia, France, and the United Kingdom. Razak et al. noted that Canada outperformed many high-income peer countries on vaccination (26). We have also suggested that cultural differences between countries, including differences in social capital and trust in government, may be important (27).

While Canada’s pandemic response, as reflected in the Oxford Stringency Index, was more stringent on average than the responses in the United States, United Kingdom, and France, it was also more stringent than Australia’s, suggesting that stringency alone cannot explain differences in outcomes. Data from Aknin et al. suggest that it may not have been stringency, but the decision to aim for elimination rather than mitigation, that resulted in the low stringency and low deaths seen in countries like Australia (28). Although more aggressive pandemic control strategies have been criticized over perceived negative mental health impacts, these authors also demonstrated that the impact of excess pandemic deaths far outweighed the impact of public health interventions as a driver of negative mental health effects during the pandemic (28). That would suggest Canada’s approach, in addition to saving more lives and costs than US and UK responses, may have been more protective of population mental health. More stringent control strategies have also been criticized as resulting in greater negative economic impacts, and indeed Canada’s GDP declined by 1.6% in the first two years of the pandemic (26); however, the $43 billion Canada effectively gained by avoiding a US-style pandemic response represents over 2% of Canadian GDP (valued at around $2.1 trillion ($CDN)).

Our analysis has three key limitations; as noted above we have not attempted to capture mental health consequences of the pandemic, or their costs. Other important costs and impacts that we do not include, and which would likely further widen the gap in health and economic consequences between these peer countries, include disutility and lost earnings associated with hospitalization, long-term costs of chronic disease, including cardiac, respiratory and neurological disease, in those who survive SARS-CoV-2 infection, and the health, economic and societal impacts of parental loss due to the pandemic (29-32). A second limitation of our analysis is our use of Ontario-specific case fatalities, and hospitalization and intensive care admission risks, to estimate outcomes averted at a national level. We use these data for pragmatic reasons: they were the most complete, and granular, Canadian death data to which we had access. However, Ontario’s epidemiology is likely similar to that of Canada overall, both because of similarities in demographics and health systems across the country, and also because the population of Ontario represents approximately 40% of the Canadian population, and 35% of Canada’s COVID-19 case load, such that the Province’s epidemiology strongly influences that of Canada as a whole. Lastly, we assume that attribution of COVID-19 deaths in Canada and comparator countries occurred in a comparable manner. The best available data (based on ratios of reported COVID-19 mortality to all-cause excess mortality during the pandemic) suggest that this is likely to have been the case for Canada, the United States, and France; reporting of COVID-19 mortality may have been more accurate in the United Kingdom than in Canada, which would tend to exaggerate the differences in outcomes between these two countries. More accurate reporting of COVID-19 deaths in Australia would lead us to underestimate the degree to which this country outperformed comparator countries (33).

In summary, we find that Canada’s relatively strong pandemic response during the first two years of the SARS-CoV-2 pandemic resulted in large numbers of deaths, hospitalizations and intensive care unit admissions averted relative to responses in the United States and United Kingdom, with more modest gains relative to France. A disease control stance focussed on elimination rather than mitigation, as was pursued in Australia during the same time period, would have resulted in further health and economic benefits.

## Data Availability

All data used for analyses are publicly available. A link to data files is included in the manuscript.

https://figshare.com/articles/dataset/Estimated_Deaths_Intensive_Care_Admissions_and_Hospitalizations_Averted_in_Canada_during_the_COVID-19_Pandemic/14036549

---

**Supplementary Table:** Mean and Standard Deviation for Oxford Pandemic Stringency Index in Canada and Comparator Countries, March 1, 2020 to May 1, 2022.

| Country | Mean | SD | P-value |
| --- | --- | --- | --- |
| Canada | 58.60 | 21.71 |  |
| Australia | 54.88 | 18.76 | <0.001 |
| France | 48.84 | 21.08 | <0.001 |
| United Kingdom | 51.14 | 24.05 | <0.001 |
| United States | 53.12 | 17.98 | <0.001 |

**NOTE:** SD, standard deviation.
P-value for Wilcoxon rank-sum test for comparison with Canada.

**Supplementary Figures:**
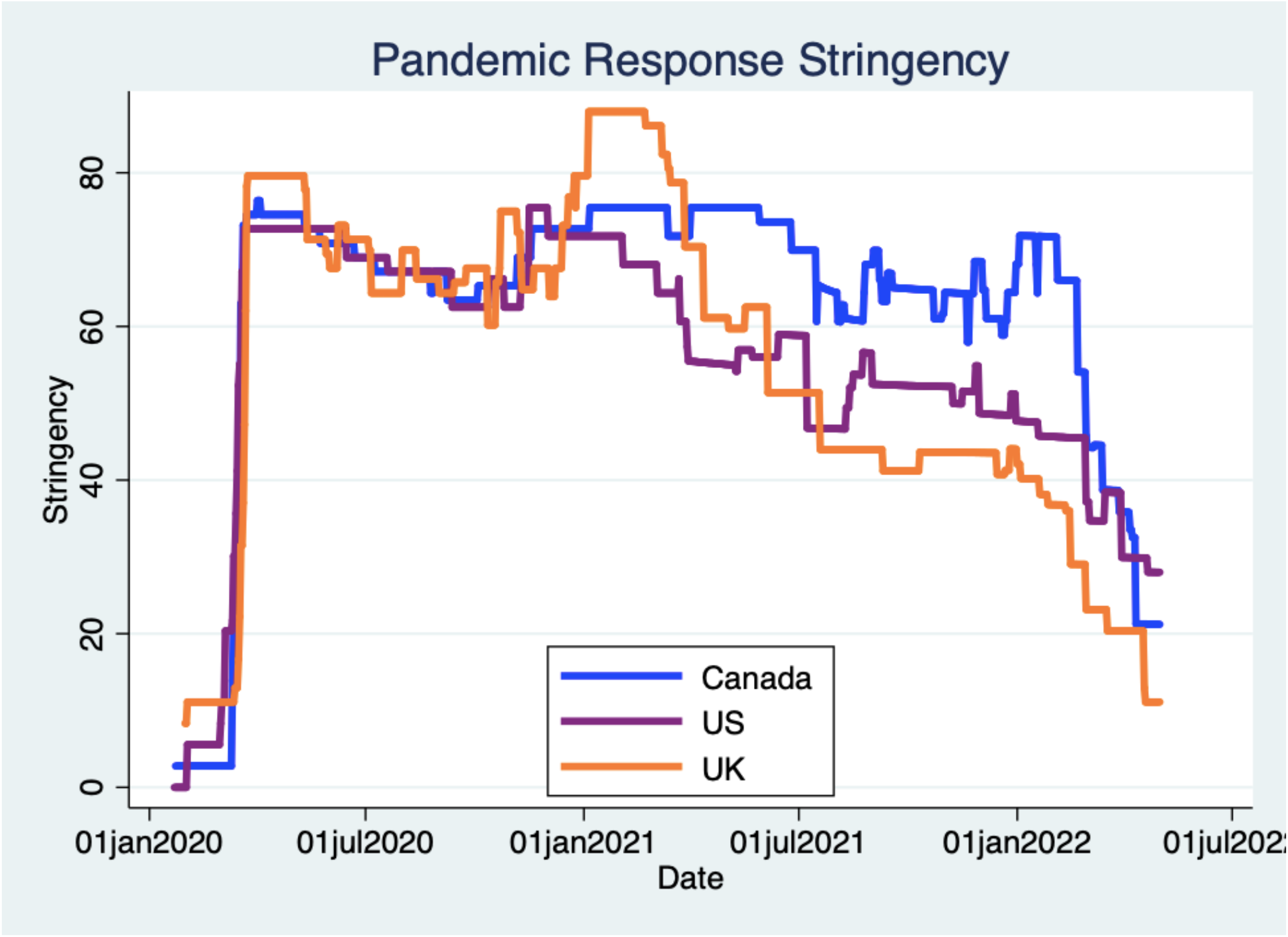
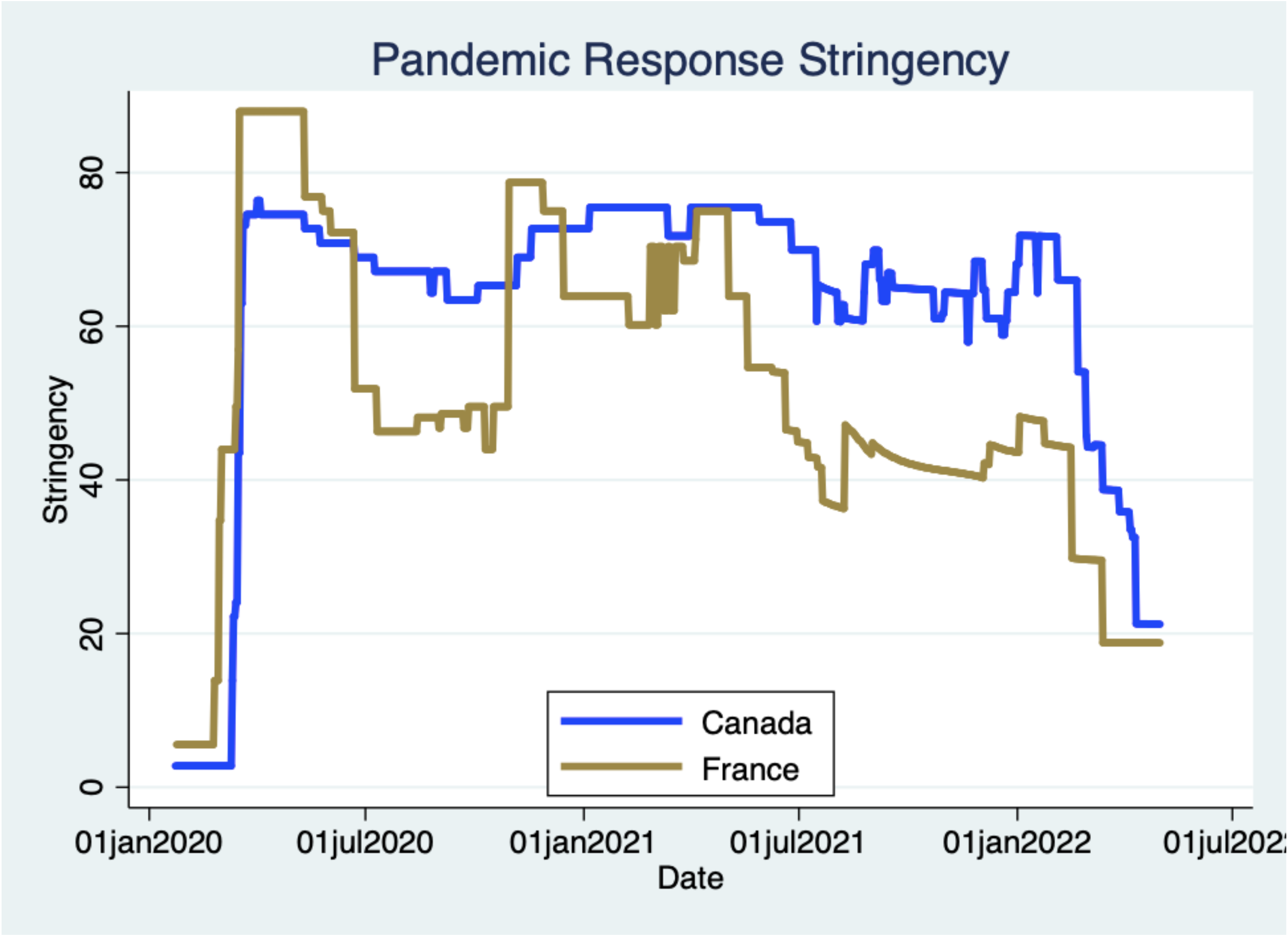
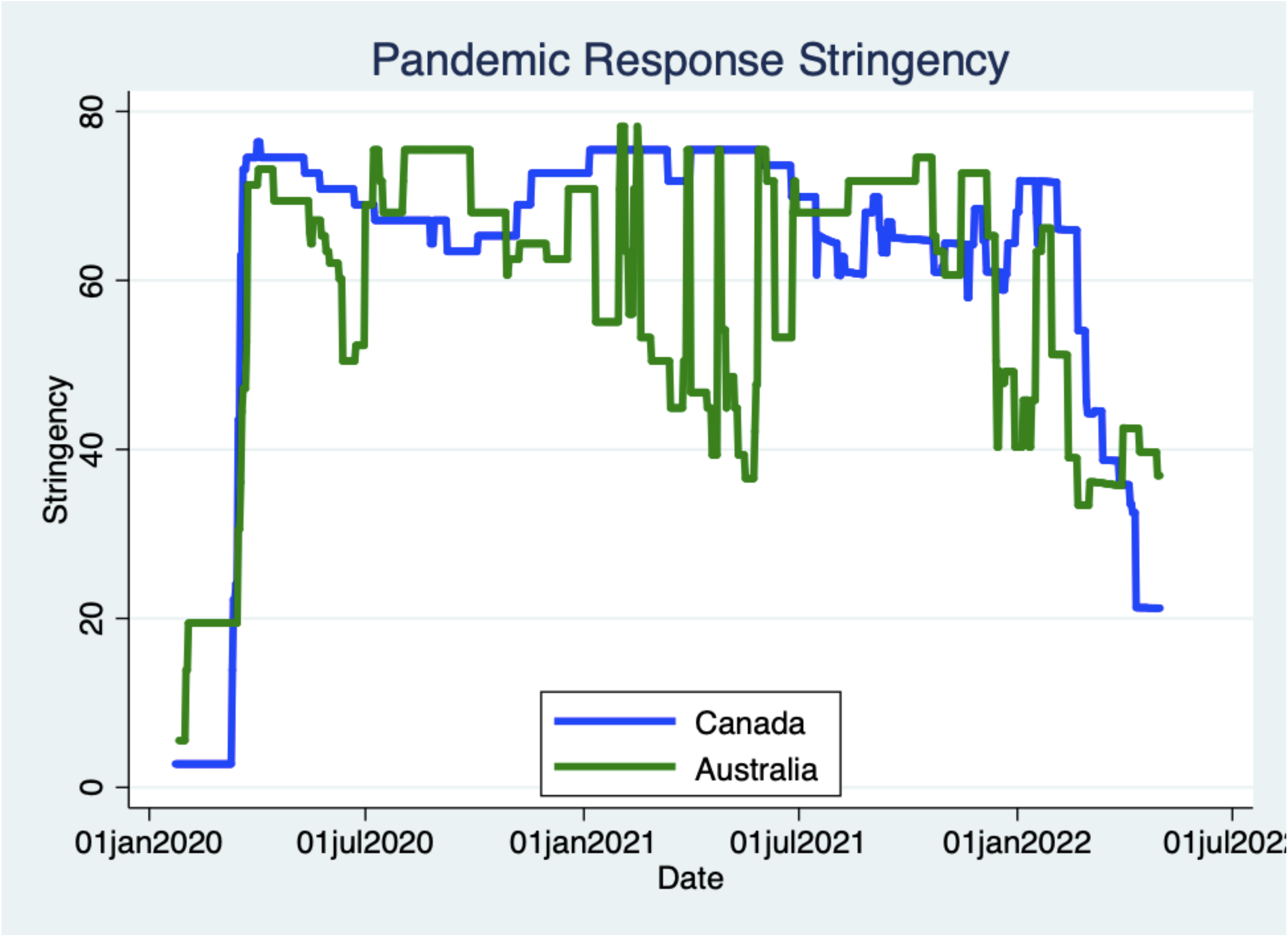
Oxford Pandemic Stringency Index by Date, Canada and Comparator Countries. Stringency values plotted to May 1, 2022. Higher values indicate more stringent control measures.

